# A Czech national administrative real-world study of diagnostics and treatment pathways of non-small-cell lung cancer stratified by disease stage: From data to actionable indicators

**DOI:** 10.64898/2026.02.20.26346704

**Authors:** Gleb Donin, Aleš Tichopád, Vratislav Sedlák, Marian Rybář, Martin Rožánek, Karla Mothejlová, Vladimír Koblížek, Pavel Turčáni, Milan Sova, Ladislav Dušek, Zuzana Bielčiková

**Author notes:** Correspondence: Zuzana Bielčiková, Department of Oncology, First Faculty of Medicine, Charles University, and General University Hospital, Prague, Czech Republic, Kateřinská 1660/32, 121 08 Prague, Czech Republic.

## Abstract

**Introduction:** Building on our previously published methodology for claims-based pathway mapping, we extended the analysis by incorporating disease staging. The aim of this study was to develop and evaluate quality indicators (QIs) in patients with non-small cell lung cancer (NSCLC).

**Methods:** This retrospective, longitudinal cohort study spanned 2017–2023, with follow-up data extending to September 2025. Data were obtained from the National Cancer Registry (NCR), the National Registry of Reimbursed Health Services (NRRHS), which is organized through seven health insurance funds providing nationwide coverage. The index date was defined as the date of the first biopsy (BX) followed by a histopathological examination (HP), along with the ICD-10 code C34. Incident patients aged ≥18 years were included if no prior malignancy was reported, and the presence of PET/CT or CT examination was mandatory in the final verified cohort. The presence of multidisciplinary team (MDT) discussion, time to treatment, availability of care in a Complex Oncology Center (COC), and completeness of predictive biomarker testing were considered key QIs.

**Results:** We analyzed the care pathways of 15,886 patients with NSCLC; 3,380 (21.3%) were not treated, and 1,837 (11.6%) were excluded due to the absence of (PET) CT prior to biopsy (BX). The final verified cohort included 10,669 patients with a median age of 69 years (interquartile range, 64–74). The incident stage distribution comprised of stage I/II (27.6%), stage III/IV (67.9%), and 4.5% unknown. Multidisciplinary team (MDT) review was reported in 53.9% of patients, with a median time to MDT discussion of 37 days. Surgery (SX) was performed in 81.0% of stage I and 68.4% of stage II patients. Fewer than 50% of patients initiated treatment within 8 weeks, regardless of disease stage. Centralization of care in COCs and implementation of MDT review showed a positive temporal trend, although disparities across disease stages and regions persisted. PD-L1 testing was documented in 70.0% of stage IV and 65.2% of stage III patients.

**Conclusions:** Administrative claims data linked with the NCR enabled stage-stratified monitoring of NSCLC care pathways and the identification of actionable QIs, which were implemented as a national tool for continuous quality evaluation of cancer care in the Czech Republic.

**KEY MESSAGES:** *What is already known on this topic:*

- Patient pathway monitoring and quality indicators (QIs) for lung cancer care — including timeliness of treatment, multidisciplinary team discussion (MDT), and centralization in specialized centers (COCs) — have been established in several European countries.
- Population-level data integrating administrative claims with cancer registry staging data to evaluate QIs across disease stages remain limited.

*What this study adds:*

- Stage-stratified analysis of 10,669 NSCLC patients revealed that fewer than 50% initiated treatment within 8 weeks, with a declining trend over time despite improvements in MDT utilization and care centralization in COCs.
- PD-L1 testing rates in stages III–IV increased over 2021–2023 but showed substantial regional variability, highlighting opportunities for improving equity of access to biomarker-guided therapy.

*How this study might affect research, practice or policy:*

- The methodology has been implemented as a national tool for continuous quality evaluation of cancer care in the Czech Republic, with PD-L1 testing completeness proposed as an additional OI alongside MDT discussion, time to treatment, and COC centralization.

## INTRODUCTION

Lung cancer is the leading cause of cancer-related mortality worldwide [1], with approximately 85% of cases classified as non-small cell lung cancer (NSCLC) [2]. Recent comprehensive evaluations of lung cancer survival in Europe report a five-year survival rate of 13%, with marked regional disparities [3]. Population-based data on lung cancer show that more than 65% of new cases in the Czech Republic are diagnosed at clinical stage III or IV [4], and similar trends are observed globally [5,6].

Monitoring patient pathways represents an important tool for improving the quality of care. A systematic review by Chiew et al. identified 56 publications on this topic from the period 2011–2019 [7]. Most of these studies focus on patient pathways [8, 9], particularly the integration of fast-track diagnostic and treatment pathways for NSCLC, and on the development and application of quality indicators (QIs) [7, 10–13], often derived from administrative or registry data, to evaluate timeliness, regional variation, and adherence to standards of care. A meta-analysis published in 2020 reported that delays in curative treatment were associated with an increased risk of death across seven major tumor types, including NSCLC [14].

Multidisciplinary team (MDT) care in NSCLC has been shown to improve coordination of diagnostic and therapeutic pathways, ensure adherence to evidence-based guidelines, and enhance patient outcomes [15–21]. Centralization of care in high-volume centers, such as in the Czech Republic within Complex Oncology Centers (COCs), further supports timely access to surgery and systemic therapy but is challenged by regional variability, reporting practices, and capacity constraints [17, 22–24].

Across Europe, QIs for lung cancer have already been implemented into routine practice in several countries including the Dutch Lung Cancer Audit (DLCA) [25], the UK National Lung Cancer Audit (NLCA) [26], and the German system of certified lung cancer centers [27]. Denmark, Norway, and Sweden routinely collect QIs through national registries and standardized cancer patient pathways [28–30], while Belgium, Italy, and Spain have developed national or regional QIs, often based on registry or administrative data [12, 31, 32]. The most frequently reported QIs include timeliness of care, MDT discussion, treatment in high-volume or certified centers, adherence to guideline-concordant therapy (e.g., surgery for stage I–II, concurrent chemoradiotherapy for stage III, molecular testing in advanced disease), and outcome measures such as 30-day postoperative survival. In addition, predictive biomarker testing is an essential prerequisite for precision therapy and a modern quality indicator in advanced NSCLC. [33].

Building on our previous claims-based methodology [34], this study integrates National Cancer Registry (NCR) data to enable stage-stratified analysis of QIs for 2017–2023. We focus on four key QI domains: MDT discussion, time to treatment, care centralization in COCs, and predictive biomarker testing completeness.

## METHODS

### Study design setting

This retrospective, longitudinal cohort study was based on routinely collected administrative health data from the Czech Republic. The Czech healthcare system provides universal coverage through mandatory enrollment in one of seven nationwide health insurance funds. Data were drawn from the National Health Information System (NHIS) and processed following a previously established methodology [34], extended in the present study by the incorporation of disease staging from the National Cancer Registry (NCR).

### Data sources

The following NHIS data sources were linked using an encrypted national patient identifier: (i) the NCR, which provided incident cancer diagnoses, ICD-O-3 morphology codes, TNM classification, and clinical stage; (ii) the National Registry of Reimbursed Health Services (NRRHS), which provided healthcare procedure claims with procedure codes, dates, medical specialty, and associated ICD-10 diagnoses; (iii) NRRHS pharmaceutical claims with Anatomical Therapeutic Chemical (ATC) classification codes; (iv) NRRHS hospitalization records; and (v) the national death data providing dates and causes of death.

### Study population and eligibility criteria

The study design and cohort selection are shown in Figure 1. The source population comprised all malignant tumors with a primary diagnosis of lung cancer (ICD-10 code C34) registered in the NCR between January 1, 2017, and December 31, 2023. Only NSCLC was included, based on ICD-O-3 morphology codes. Patients were excluded if they met any of the following criteria: age younger than 18 years at diagnosis; a prior malignancy of any type recorded in the NCR (excluding non-melanoma skin cancer, ICD-10 C44); a non-first C34 diagnosis in the NCR (i.e., second or subsequent lung tumor); absence of any administrative claims record; discrepancy in birth year exceeding one year or in date of death exceeding seven days between the NCR and NRRHS; death occurring at or before the date of diagnosis; absence of a valid index date (see below); cancer treatment documented before the index date; discrepancy between the index date and the NCR diagnosis date exceeding 365 days; or index date falling outside the study period 2017–2023.

**Figure 1.**
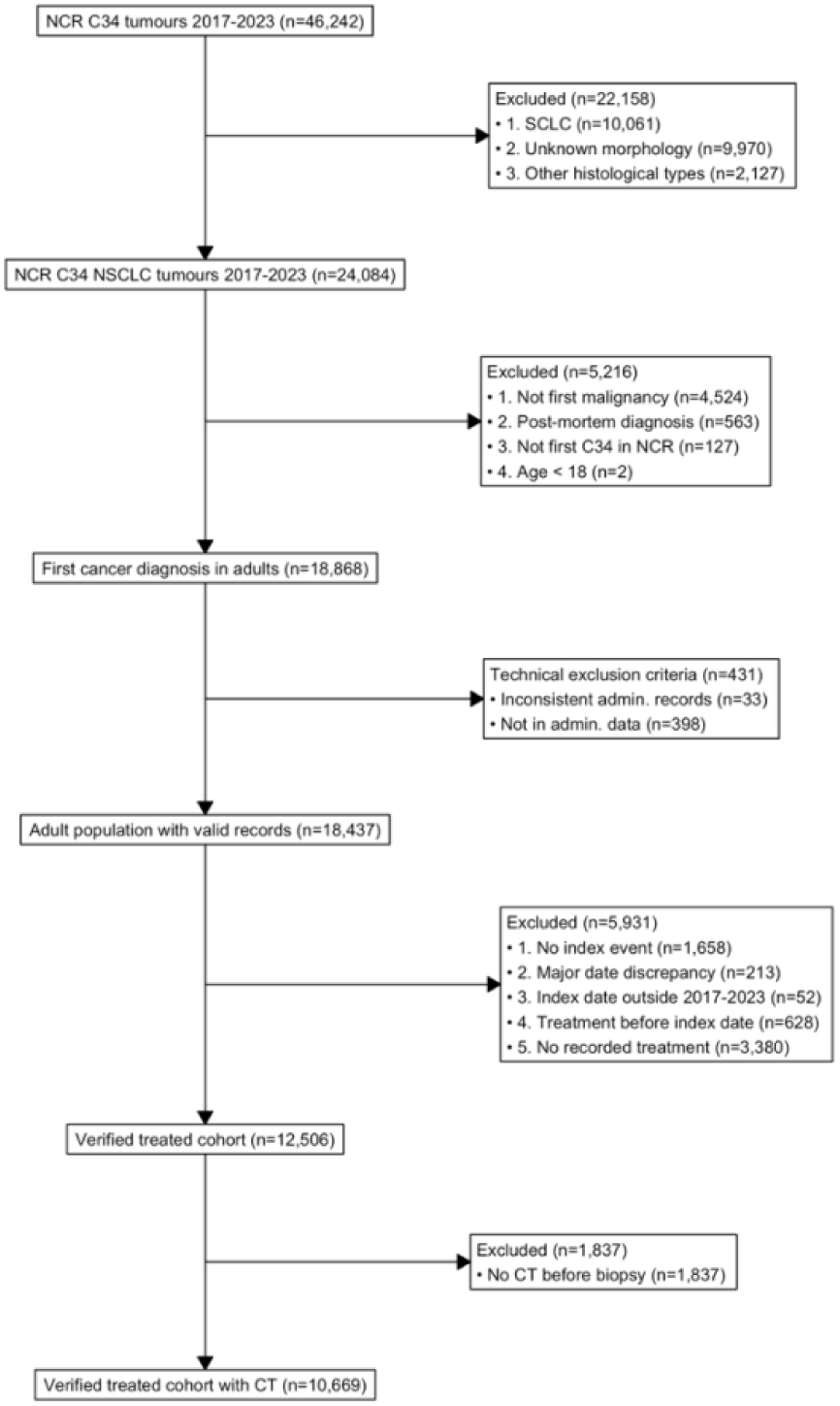
Overview of study design and patient cohort selection *Abbreviations: NCR National Cancer Registry, ICD-10 International Classification of Diseases, 10^th^ revision; C34 lung cancer, CT Computed tomography*

Among patients meeting all eligibility criteria (the analytical cohort), those with a documented first-line treatment (FLT) constituted the treated cohort. The treated cohort was further refined by requiring evidence of computed tomography (CT, including PET/CT) imaging within 365 days before the index biopsy, yielding the verified treatment cohort with CT. This criterion was applied to ensure diagnostic plausibility, as treatment initiation without prior cross-sectional imaging is inconsistent with standard-of-care practice. Time to treatment, time to MDT, and other time-to-event intervals were calculated from the date of the last CT or PET/CT examination preceding the biopsy, unless stated otherwise.

### Index date

The index date was defined as the date of the first lung biopsy (BX) followed by a histopathological examination (HP) within 60 days, with confirmation of the ICD-10 diagnosis code C34. Biopsy procedures included bronchoscopic techniques (standard needle aspiration and endobronchial ultrasound-guided biopsy) as well as other lung or pleural tissue sampling procedures identified by national procedure codes. Diagnostic confirmation required that code C34 be reported on a claim associated with the BX or the HP, or on any claim submitted within 180 days following the HP. When multiple valid BX–HP pairs existed for a patient, the earliest biopsy date was selected.

### Clinical event classification

All outpatient and inpatient claims were mapped to standardized clinical event types using predefined algorithms based on procedure codes, ATC codes, and ICD-10 diagnosis filters. The following event categories were defined: (i) diagnostic, including BX, HP, and CT (PET/CT) imaging, (ii) therapeutic, including surgery (SX), radiotherapy (RT), and pharmacotherapy (PHT), with subtypes for chemotherapy (PHT_CT), targeted therapy (PHT_TT), and immunotherapy (PHT_IO), and (iii) MDT and predictive biomarker testing.

### Treatment definitions

FLT was defined as the first occurrence of SX, RT, or PHT after index date. When multiple treatment types were recorded on the same date, event type priority was applied (SX > PHT > RT). Within PHT, we distinguished between PHT_CT, including platinum compounds, antimetabolites, taxanes, and alkylating agents; PHT_TT, including EGFR, ALK, BRAF, MET, RET, and ROS1 inhibitors, as well as antiangiogenic agents; and PHT_IO, including PD-1, PD-L1, and CTLA-4 checkpoint inhibitors. Neoadjuvant chemotherapy (PHT_NEO) was defined as any chemotherapy administered prior to SX and followed by SX within 12 months of its initiation. Treatment activity within 12 months after FLT was used to characterize the main treatment modality.

### Quality indicators

Four key QIs were assessed. First, MDT was identified using a dedicated procedure code for tumor board review and measured as the first occurrence between the index date and FLT. Second, time to treatment was defined as the number of days from the last CT or PET/CT prior to BX to FLT initiation, and was categorized using cut-offs of 4, 6, and 8 weeks. Third, centralization of care was assessed by matching the facility where FLT was delivered to a national list of COCs. Fourth, completeness of predictive biomarker testing was evaluated as described below.

### Predictive biomarker testing

Predictive biomarker testing was identified from claims data using national procedure codes for molecular diagnostic techniques. Prior to 2021, only non-specific procedure codes were available for molecular diagnostics (quantitative PCR, fluorescence in situ hybridization, immunohistochemistry, and next-generation sequencing), precluding the identification of individual biomarkers. Beginning in 2021, dedicated procedure codes were introduced for each biomarker, enabling biomarker-specific analyses. Consequently, PD-L1 testing analyses were restricted to 2021–2023. The PD-L1 testing rate was defined as the proportion of patients with a documented PD-L1 testing event.

### Follow-up and censoring

The follow-up period was defined as the time from the index date to the earliest of the following events: (i) death (as recorded in the national death registry), (ii) the date of the last available claims record for the patient, or (iii) the administrative end of data availability (September 30, 2025). Survival time was calculated as the number of days from the index date to the censoring event. Patients who had not died by the end of follow-up were censored.

### Statistical analysis

Descriptive statistics were reported as absolute and relative frequencies for categorical variables and as medians with interquartile ranges (IQR) for continuous variables. Time to treatment was analyzed by type of FLT, disease stage, calendar year, and region of residence. MDT utilization, COC centralization, and predictive biomarker testing rates were analyzed by disease stage, calendar year, and region. Treatment modalities were compared across disease stages and calendar years using stacked bar charts and tabulated proportions. The statistical analysis was conducted using R version 4.4 [35].

## RESULTS

### Study cohort

The analytical cohort comprised 15,886 NSCLC patients who met all eligibility criteria (Figure 1). Of these, 78.7% (12,506) received FLT (treated cohort) and 21.3% (3,380) did not. Among treated patients, 14.7% (1,837) were excluded due to the absence of CT or PET/CT prior to BX. The verified treatment cohort with CT included 10,669 patients, comprising 62.7% (6,690) males and 37.3% (3,979) females. The median age at index was 69 years (interquartile range, 64–74). The distribution of patients across disease stages was as follows: 17.0% (1,811) in stage I, 10.6% (1,132) in stage II, 27.2% (2,901) in stage III, 40.7% (4,342) in stage IV, 0.9% (101) in stage X (unknown stage due to objective reasons, e.g., poor clinical condition or terminally ill patient), and 3.6% (382) in stage Y (unknown stage due to unspecified reasons).

The untreated group (N = 3,380; 21.3%) comprised 67.7% males (2,288) and 32.3% females (1,092), with a median age of 72 years (interquartile range, 66–77). Stage distribution was as follows: 3.0% (100) in stage I, 2.9% (98) in stage II, 17.0% (573) in stage III, 65.5% (2,213) in stage IV, 7.2% (244) in stage X, and 4.5% (152) in stage Y. The median follow-up was 1.6 months (interquartile range, 0.8–3.5). By the end of the study period, 97.4% (3,291) of untreated patients had died, with a median time to death of 1.6 months.

### Patient pathways and MDT

For the majority of patients, discrepancies in timing were observed between the index date (BX) and the date of diagnosis recorded in the NCR (see supplemental Table S1).

Patient pathways in the verified treatment cohort with CT are presented online (see Supplemental Figures S1A). Following the diagnosis of NSCLC, 53.9% (5,750) of patients received an MDT review within a median of 37 days (interquartile range, 23–61). The indication for MDT review showed a positive trend over time; however, differences were observed between clinical stages as well as across regions (online supplemental Figures S2A–B). MDT utilization increased over time, rising from 43–45% in 2017 to 69–74% in 2023 in early stages I–II, from 42% to 73% in stage III, and from 37% to 58% in stage IV.

### Treatment modalities

In the overall population of treated patients, the FLT modalities were distributed as follows: SX in 29.5% (3,150) of patients, RT in 15.7% (1,679), and PHT in 54.7% (5,840) (Supplemental Figures S1A). With respect to disease stage, SX was performed in 81.0% (1,467) of stage I patients and 68.3% (774) of stage II patients, whereas PHT_CT was administered to 55.5% (1,610) of stage III patients and 48.3% (2,100) of stage IV patients (online supplemental Figures S1B–E).

Figure 2A illustrates the main treatment modalities including SX in early stages, and PHT in advanced stages of NSCLC patients. In stages I and II, a decreasing trend in SX and an increasing trend in RT were observed over time, while in stage IV the proportion of patients treated with PHT increased. Figure 2B provides a detailed summary of FLT. The key findings include the increasing trend in (probably stereotactic) RT in stage I, the increasing trend in neoadjuvant therapy including PHT_IO in stage II, and the increasing availability of PHT_IO/PHT_TT in advanced stages.

**Figure 2.**
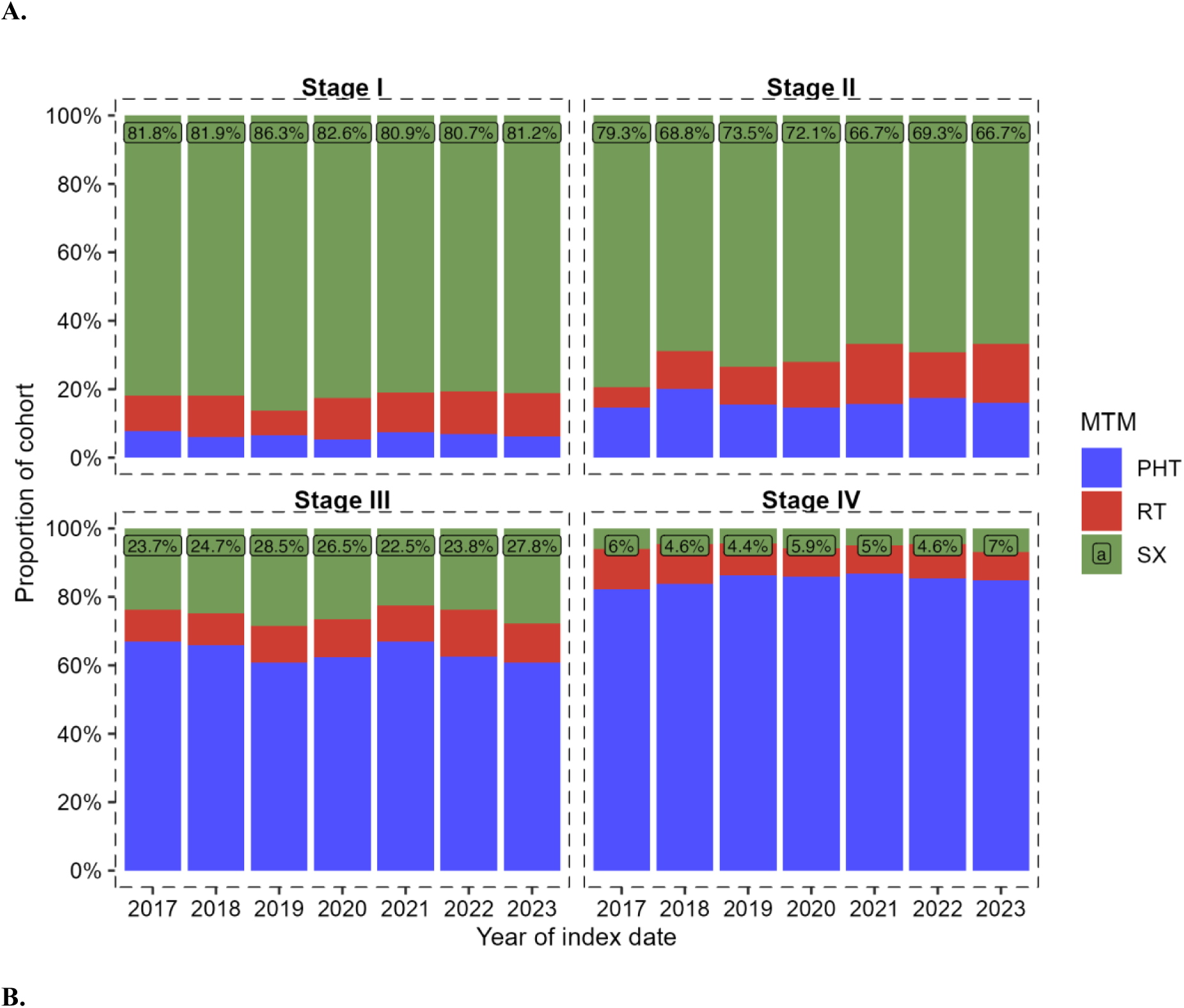
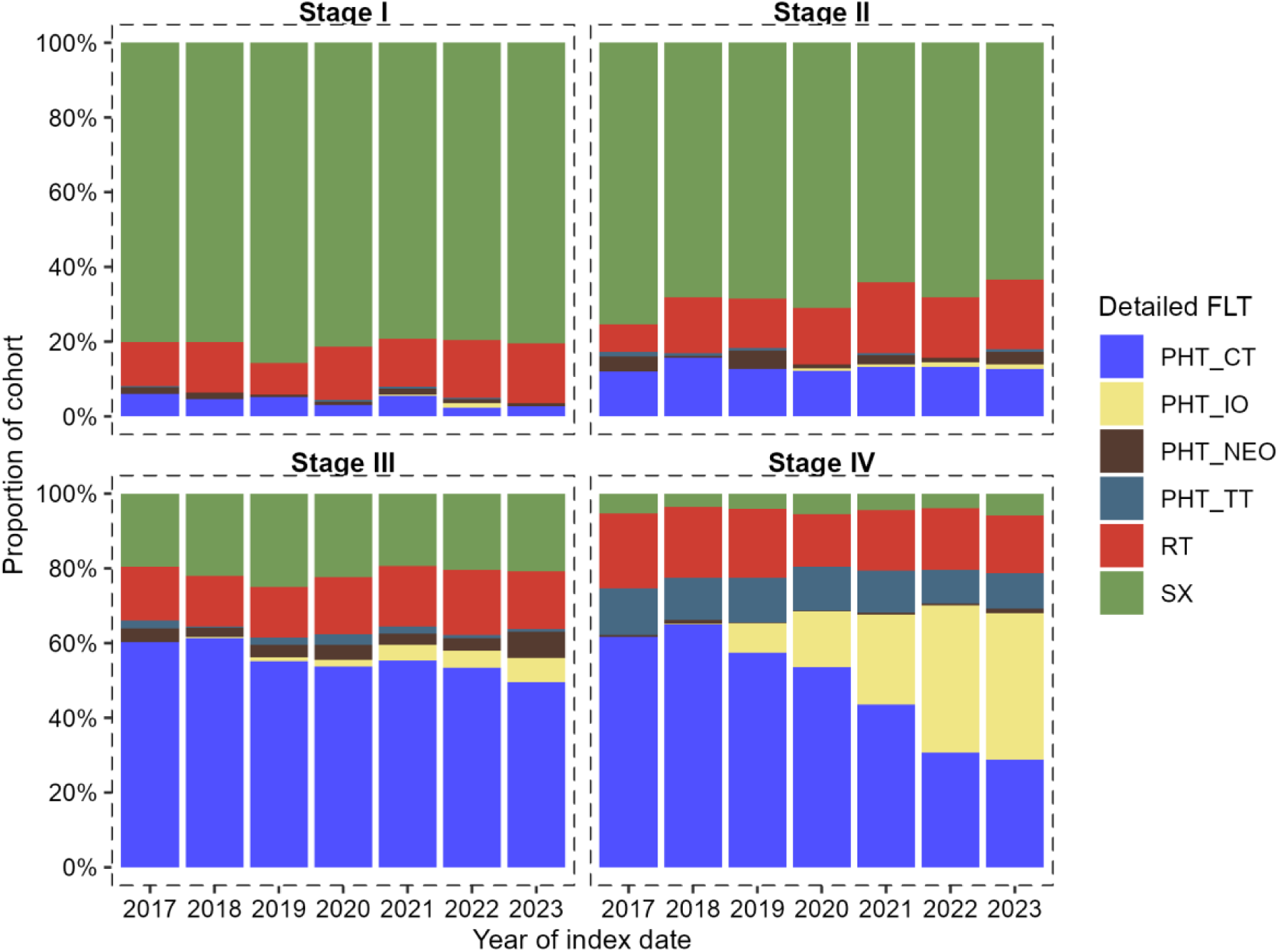
Treatment modalities by disease stage across time A. The definition of the main treatment modality (MTM) included cases with a sequence of two treatment methods occurring within a 24-week time window. This primarily involved patients indicated for sequential pharmacotherapy (PHT) and radiotherapy (RT) or those who received neoadjuvant therapy followed by surgery (SX). B. Detailed first-line therapy by disease stage across time *Abbreviations: PHT pharmacotherapy, CT chemotherapy, IO immunotherapy, NEO neoadjuvant chemotherapy, TT targeted therapy, RT radiotherapy, SX surgery*

### Predictive biomarker testing

PD-L1 testing was analyzed for the period 2021–2023, following the introduction of dedicated procedure codes that enabled identification of specific biomarkers from claims data. PD-L1 testing was documented in 55.5% (2021), 60.5% (2022), and 61.0% (2023) of treated patients across stages I–IV (Figure 3). PD-L1 testing rates were strongly dependent on disease stage. In stages III and IV, PD-L1 testing rates reached 65.2% and 70.0% in 2023, respectively. In early stages I and II, PD-L1 testing rates were lower (39.6% and 50.7% in 2023), consistent with the predominance of surgical management. Baseline characteristics of PD-L1-tested and non-tested patients in stages III–IV during 2021–2023 are provided in online supplemental Table S2.

**Figure 3.**
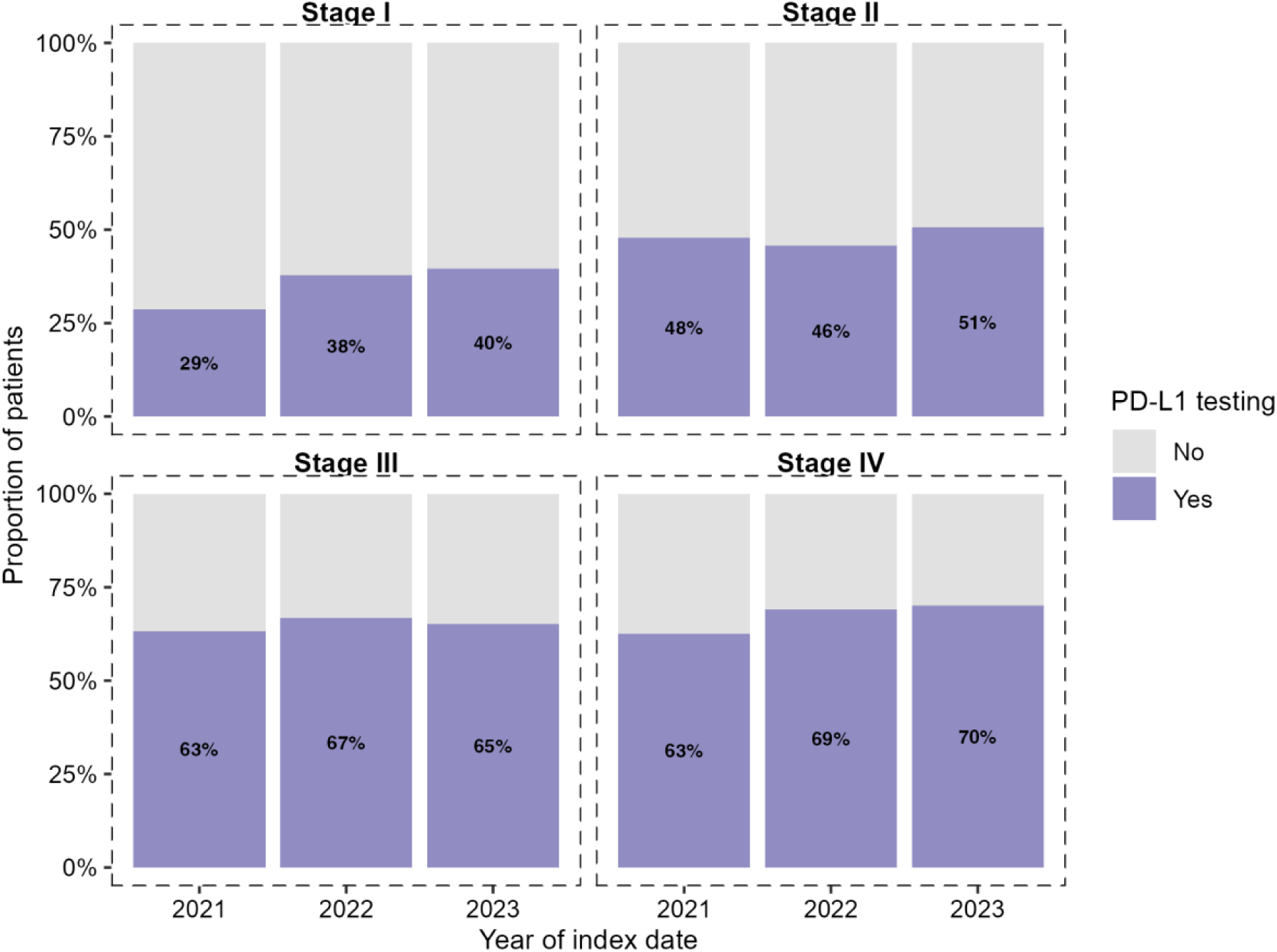
PD-L1 testing by disease stage and year (2021-2023)

### Time to treatment

The median time to treatment initiation measured from (PET) CT depended on the type of FLT and disease stage, reaching 64–71 days in patients treated with SX, 55–72 days in patients treated with PHT, and 45–118 days in patients treated with RT (see online supplemental Table S3A).

The median time to treatment was compared across time periods and disease stages (Figure 4). A 4-week cut-off was met in approximately 5–10% of patients in stages I-III disease and in 15% of patients with stage IV disease. An 8-week cut-off was achieved in fewer than 50% of patients with stages I–II disease and in 40–50% of patients with stages III–IV. Further details are provided online (in supplemental Table S3B).

**Figure 4.**
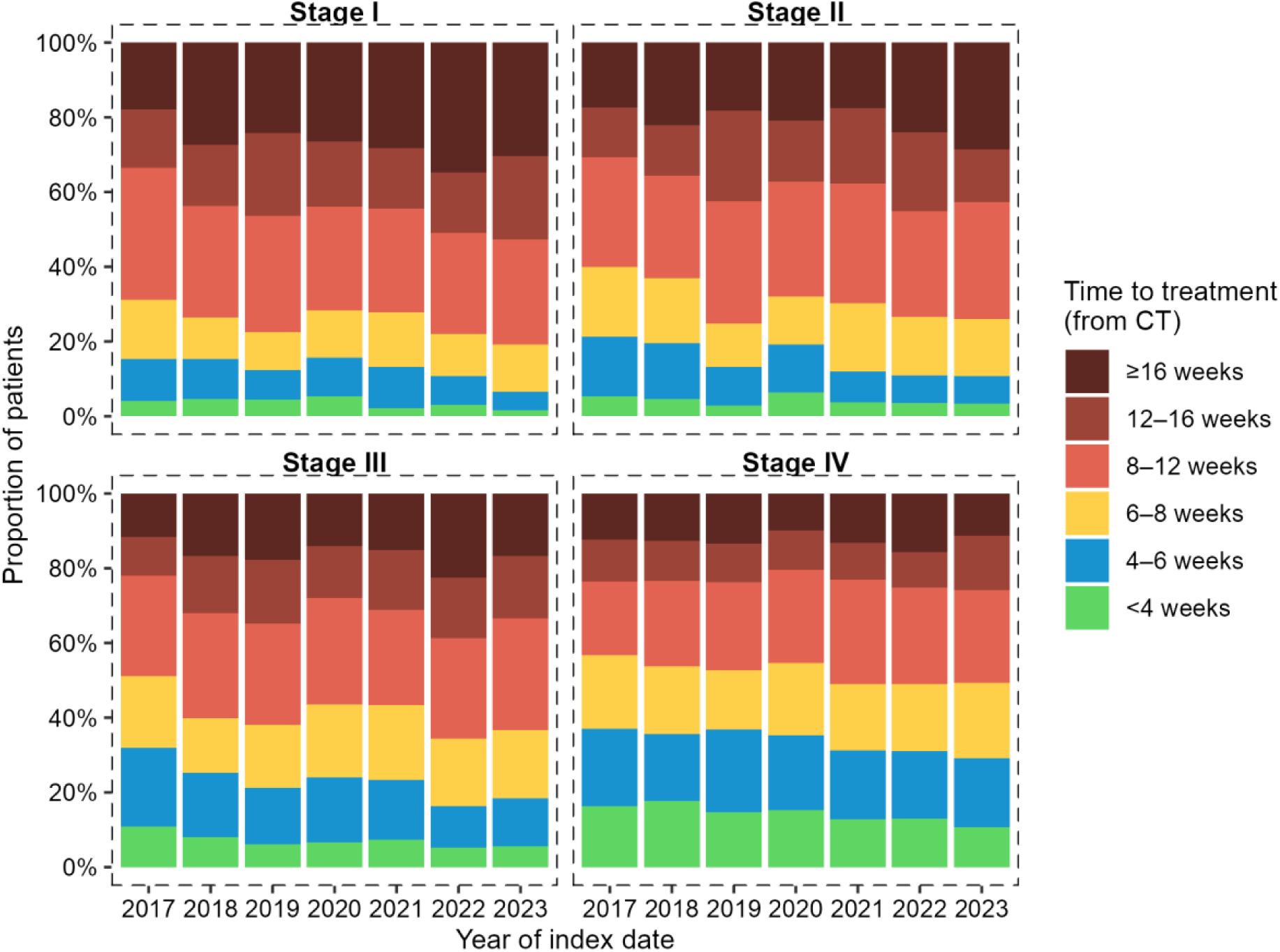
Time from (PET) CT to treatment initiation by disease stage across time

When the time to treatment was divided into the interval from (PET) CT to MDT discussion and from MDT to FLT, it became evident that the main delay was associated with referral of the patient to the MDT. In contrast, a marked acceleration in time to treatment was observed in the subsequent phase (see online supplemental Figures S3A-B). Time to treatment also varied according to the patient’s region of residence (see online supplemental Figure S4).

### Treatment within COC

Care centralization in the Czech Republic has increased over time. In stages I and II, almost all patients received FLT in COCs. In stages III and IV, the growing proportion of patients treated in COCs is particularly evident (Figure 5).

**Figure 5.**
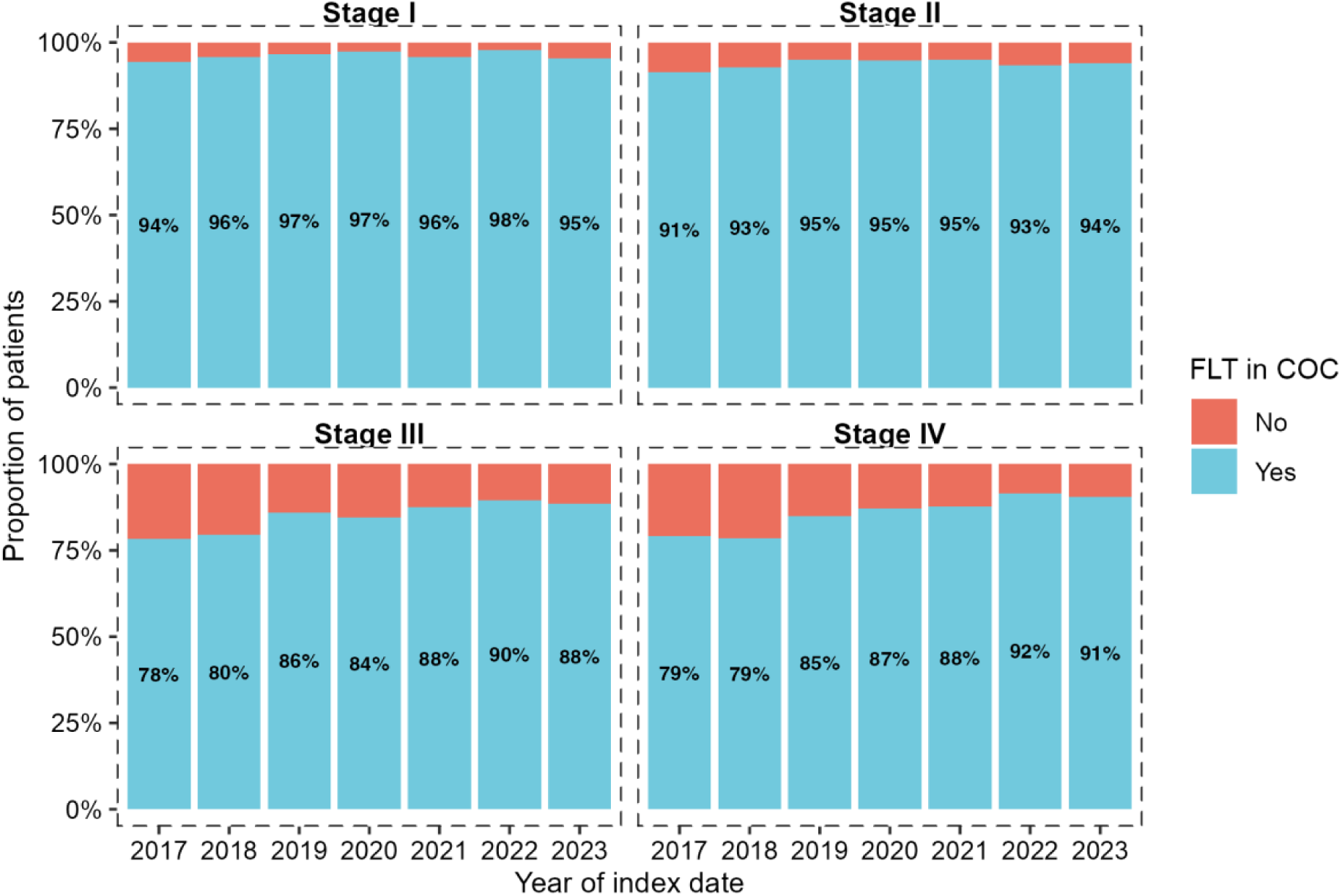
First-line therapy (FLT) in Complex Oncology Centres (COCs) among the verified treated cohort with CT across time

Nevertheless, substantial regional variability was observed across the Czech Republic. In 2023, the proportion of patients treated in COCs ranged from 80% to 100% in stage I, from 75% to 100% in stage II, from 60% to 100% in stage III, and from 71% to 100% in stage IV, depending on the patient’s region of residence (online supplemental Figure S5).

## DISCUSSION

NSCLC is a fast-track diagnosis in which prognosis can be strongly influenced by early access to treatment. These characteristics make NSCLC a suitable model for demonstrating the importance of systematic patient pathway monitoring. We have developed a comprehensive model that links data from the NCR, NRRHS, and administrative payer records with the aim of mapping patient pathways across different diagnoses and integrating them into the process of evaluating the quality of care provided. Compared with our previous study [34], which analyzed 5,819 patients during 2018–2021 using only administrative claims data without staging information, the present analysis integrates NCR data and extends the cohort to 15,886 patients diagnosed during 2017–2023. This extension enabled stage-stratified analysis of all QIs and provided a more complete picture of NSCLC care in the Czech Republic. Both clinical staging and expert feedback are reflected in the presented results.

Mapping real-world care trajectories allows the identification of delays and organizational barriers, thereby supporting targeted improvements in the quality and accessibility of cancer care [7–13]. In this study, we described individual points along the patient pathway that can be measured across different regions of the Czech Republic and over time, serving as effective QIs for assessing access to care. Region-specific QIs describing the catchment areas of COCs, as well as local QIs reflecting COC activities – such as the proportion of MDT consultations by diagnosis, time from first patient contact with the COC to treatment initiation, or the proportion of patients receiving center-based therapy – are already embedded in national legislation. Numerous examples across Europe demonstrate that QIs are increasingly used not only to assess clinical practice but also to reduce regional variation and drive improvements in lung cancer care [26–33].

Administrative data play a crucial role in refining patient pathway information, as registry data alone may be incomplete or inaccurate. By applying an administratively defined index date, our model produces a streamlined verified cohort, enabling pathway mapping for approximately half of all NSCLC patients diagnosed and treated in the Czech Republic between 2017 and 2023. A major remaining challenge is shifting the index date earlier in the disease course, ideally enabling detection of NSCLC in the early phase. Recent studies have explored computational approaches applied to electronic health records and primary care documentation as a means of facilitating earlier identification of lung cancer [36–38].

Disease stage emerged as a key determinant in the interpretation of our data. Fewer than 5% of patients had an unknown stage, supporting the robustness of the analysis. The proportion of untreated patients (21.3%), their predominance of advanced-stage disease, very short median survival, and high mortality rate suggest that the vast majority had a performance status precluding active treatment. Nevertheless, the proportion of untreated cases in stages I and II (3.0% and 2.9% of untreated patients, corresponding to approximately 200 individuals with potentially curable disease) appears high given current therapeutic options. Possible explanations include severe comorbidities or poor performance status precluding treatment, patient refusal, or delays in referral leading to clinical deterioration before treatment initiation. These cases warrant targeted investigation, as they may represent missed opportunities for curative intervention. For the same reason, some patients may not be referred to a COC for MDT evaluation and may instead be managed outside the COC network.

Regarding diagnostics, 14.7% of patients were excluded from the final verified cohort due to the absence of a (PET) CT prior to biopsy, raising concerns about the quality of the initial diagnostic work-up. Treatment initiation in patients whose only prior examination was an X-ray is unrealistic and was therefore excluded. The absence of CT imaging may reflect regional differences in waiting times or other system-related factors.

With respect to treatment patterns, the observed slight decline in SX for stages I–II disease was unexpected and may reflect the growing adoption of stereotactic body radiotherapy (SBRT), which has emerged as a guideline-recommended alternative for medically inoperable early-stage NSCLC [39]. In stage II, the increasing proportion of neoadjuvant therapy is consistent with evolving evidence supporting perioperative chemoimmunotherapy regimens [40, 41]. These treatment pattern shifts suggest that Czech clinical practice is progressively aligning with international guideline updates. The occurrence of PHT_CT as FLT in stage I is difficult to interpret and may be attributable to reporting inaccuracies. Surgery as FLT in stage IV disease was rare and likely represents metastasectomy or palliative resection.

The central focus of our analysis was the time to treatment. A recent systematic review has demonstrated substantial heterogeneity in the definition of diagnostic intervals, with starting points ranging from THE first symptom to the initial general practitioner consultation, or the first abnormal imaging finding [42]. Evidence regarding the most critical interval for treatment initiation in lung cancer remains inconsistent, with some studies using the initial suspicion of cancer as the index date [42,43]. Our analysis revealed substantial variation in time to treatment by treatment modality, disease stage, and region, with only a minority of patients meeting the recommended 4- or 8-week thresholds. Nevertheless, these findings are consistent with European data, indicating comparable treatment intervals in the Czech Republic [42].

Patients with earlier-stage disease (I–II) experienced longer intervals from CT to MDT discussion and from BX or CT to treatment initiation. Similar findings were reported by Stokstad et al., who attributed delays primarily to late referral for PET-CT and repeated biopsies following unsuccessful bronchoscopy [44]. Although we did not assess the number of biopsies, prolonged diagnostics in early-stage disease may not represent a clinical issue, as it often reflects follow-up of pulmonary nodules with a low risk of rapid progression. As illustrated in Figure 4, very rapid treatment initiation following CT was uncommon and may even correspond to SX of non-verified nodules.

The proportion of patients initiating treatment within 8 weeks declined over time across all stages, despite improvements in MDT utilization and COC centralization (Supplementary Table S2B). This trend may reflect increasing diagnostic complexity, including broader use of predictive biomarker testing, greater centralization with associated logistical delays, and potential capacity constraints at COCs. The apparent trade-off between improving process indicators and worsening timeliness warrants further investigation.

PD-L1 testing (2021–2023) showed a clear stage-dependent gradient, reaching 70.0% in stage IV and 65.2% in stage III by 2023, compared with 39.6% in stage I. This aligns with its primary role in guiding immunotherapy in advanced disease. Population-level data on biomarker testing completeness remain limited in Europe [25, 26], and our findings provide a national benchmark for future comparisons.

Although expanded biomarker testing likely contributes to longer diagnostic pathways, biomarker-guided treatment selection is expected to improve outcomes. We therefore propose PD-L1 testing completeness as an actionable QI, alongside MDT discussion, time to treatment, and COC centralization, to support ongoing monitoring of diagnostic quality in NSCLC care.

Evidence regarding the prognostic impact of surgical delay remains mixed, with some studies reporting adverse effects [45], while others found no significant association [46, 47]. In our cohort, only 5% of patients with stage I-II disease underwent SX within 4 weeks, and approximately one-third within 8 weeks. Although we did not directly assess prognostic outcomes, timely treatment is likely particularly important in stage II disease. Potential benefits of surgical centralization in 2020–2021 may have been offset by delays related to the COVID-19 pandemic.

Patient pathway monitoring and MDT organization have consistently emerged as key QIs for optimizing lung cancer care [15, 23–24]. In our study, MDT indication and centralization of care in COCs demonstrated a positive temporal trend, although substantial disparities persisted across stages and regions. Centralization was most pronounced in advanced stages, yet regional variability remained considerable — for example, in 2023, the proportion of stage IV patients treated within COCs ranged from 71% to 100% across regions. Clinical guidelines emphasize that NSCLC care should be delivered in centers with MDTs comprising thoracic surgeons, oncologists, pulmonologists, radiologists, and other specialists [48]. These findings underscore the need for further standardization of care pathways to reduce regional inequalities and ensure equitable access to comprehensive multidisciplinary care.

The main limitations of this study include its restriction to a selected cohort, with complete data available for approximately half of newly diagnosed NSCLC patients; the definition of the index date as the first biopsy rather than symptom onset or the initial chest X-ray, which may underestimate diagnostic delays; and the lack of data on the number of biopsies and patient performance status, both of which strongly influence MDT indication, centralization, and treatment selection.

In addition, reliance on procedure codes to ascertain biomarker testing may underestimate true testing rates, even after the introduction of dedicated codes in 2021. Regional differences in MDT indication and PD-L1 testing may also reflect variations in reporting practices, COC capacity, transportation logistics, or social accessibility, as illustrated by the exceptionally low documentation of PD-L1 testing in the Pilsen region, which likely reflects incomplete procedure code reporting rather than actual clinical practice.

Although the study period overlapped with the COVID-19 pandemic (2020–2021), we did not observe clear disruptions in patient volumes or time-to-treatment trends across disease stages, suggesting that lung cancer care pathways in the Czech Republic remained relatively resilient during this period. While some European studies reported modest reductions in lung cancer diagnoses and a shift toward more advanced stages during the pandemic [49], our findings suggest that the overall impact on treatment pathways in the Czech Republic was limited.

Conversely, the strengths of this study lie in the integration of administrative data with the NCR and NRRHS, providing highly specific and complementary information; the reflection of real-world diagnostic and therapeutic practice, including CT, PET/CT, surgical centralization, PD-L1 testing, and use of immunotherapy and targeted therapy; the ability to monitor regional availability of care; and the evaluation of COC performance using indicators such as MDT reporting, PD-L1 testing completeness, and time to treatment.

## CONCLUSIONS

We developed and implemented a methodology for processing administrative data as a technical infrastructure to support the organization and quality evaluation of cancer care in the Czech Republic. By integrating multiple data sources—administrative data, NRRHS, and NCR—we minimized their individual limitations. Patient pathway monitoring reflects real-world practice, reveals regional disparities, and provides a robust basis for evaluating care quality, with pathway mapping itself serving as a QI. Several pathway-related QIs, including those capturing COC activities, are already embedded in national legislation. Integrating pathway-based indicators into NSCLC management should account for clinical, economic, and logistical considerations to inform policies aimed at optimizing care delivery and improving patient outcomes.

## DECLARATIONS

### Acknowledgments

The data were kindly provided by the Health Insurance Bureau in Prague, Czech Republic.

## Funding

This research was supported by the Grant of the Operational Programme Jan Amos Komenský (OP JAK) with project AGEING-CZ No. CZ.02.01.01/00/23_025/0008743.

## Disclosure

The authors declare no competing or financial interests.

## Authors’ contributions

Conception/design: A.T., G.D., M.R. Investigation: G.D. Data analysis: A.T., G.D. Project administration: A.T. Data interpretation: A.T., G.D., Z.B., L.D., V.K., V.S., P.T. Manuscript writing: Z.B., A.T., G.D. Manuscript editing: K.M., L.D., V.K., V.S., P.T., J.M. Final approval of the manuscript: All authors.

## Ethics approval

This study was conducted using anonymized retrospective administrative claim data, and approval by an ethics committee was not required.

## Data availability

The data remains property of the insurance companies and cannot be shared without their permission.

## SUPPLEMENTARY MATERIALS

### Additional file 1 (PDF format)

**Table S1** Absolute difference between the index date (biopsy, BX) and the date of diagnosis according to the National Cancer Registry (NCR)

**Figure S1** Pathway trajectories by disease stage in the verified treatment cohort with CT

**Figure S2** Multidisciplinary team (MDT) review in the verified treated cohort with CT over time and by region

**Table S2** Baseline characteristics of PD-L1-tested and non-tested patients (stages III–IV, 2021–2023)

**Table S3** Time to treatment initiation by disease stage and type of first-line therapy (FLT)

**Figure S3** Time to MDT and from MDT to treatment initiation in verified treated cohort

**Figure S4** Time to treatment initiation by disease stage and region of residence

**Figure S5** First-line therapy (FLT) in Complex Oncology Centers (COCs) for treated patients by region of residence and disease stage

## Notes

### Competing Interest Statement

The authors have declared no competing interest.

### Funding Statement

This research was supported by the Grant of the Operational Programme Jan Amos Komensky (OP JAK) with project AGEING-CZ No. CZ.02.01.01/00/23_025/0008743.

### Author Declarations

Data were obtained from the National Cancer Registry (NCR), the National Registry of Reimbursed Health Services (NRRHS), which is organized through seven health insurance funds providing nationwide coverage. Datasets was fully de-identified.

