## supplementary_material for "A Czech national administrative real-world study of diagnostics and treatment pathways of non-small-cell lung cancer stratified by disease stage: From data to actionable indicators"

Table S1 Absolute difference between the index date (biopsy, BX) and the date of diagnosis according to the National Cancer Registry (NCR)

Absolute difference between index date (BX) and diagnosis date (NCR)

| | **Stage** | | | | | | | | | | | |
|---|---|---|---|---|---|---|---|---|---|---|---|---|
| **Difference** | **I** | | **II** | | **III** | | **IV** | | **X** | | **Y** | |
| | **N** | **%** | **N** | **%** | **N** | **%** | **N** | **%** | **N** | **%** | **N** | **%** |
| < 1 week | 482 | 26.6% | 411 | 36.3% | 1 478 | 50.9% | 2 466 | 56.8% | 57 | 56.4% | 212 | 55.5% |
| < 1 month | 952 | 52.6% | 696 | 61.5% | 2 209 | 76.1% | 3 593 | 82.7% | 82 | 81.2% | 293 | 76.7% |
| < 6 months | 1 772 | 97.8% | 1 115 | 98.5% | 2 878 | 99.2% | 4 300 | 99.0% | 100 | 99.0% | 374 | 97.9% |
| All | 1 811 | 100.0% | 1 132 | 100.0% | 2 901 | 100.0% | 4 342 | 100.0% | 101 | 100.0% | 382 | 100.0% |

Figure S1 Pathway trajectories by disease stage in the verified treatment cohort with CT

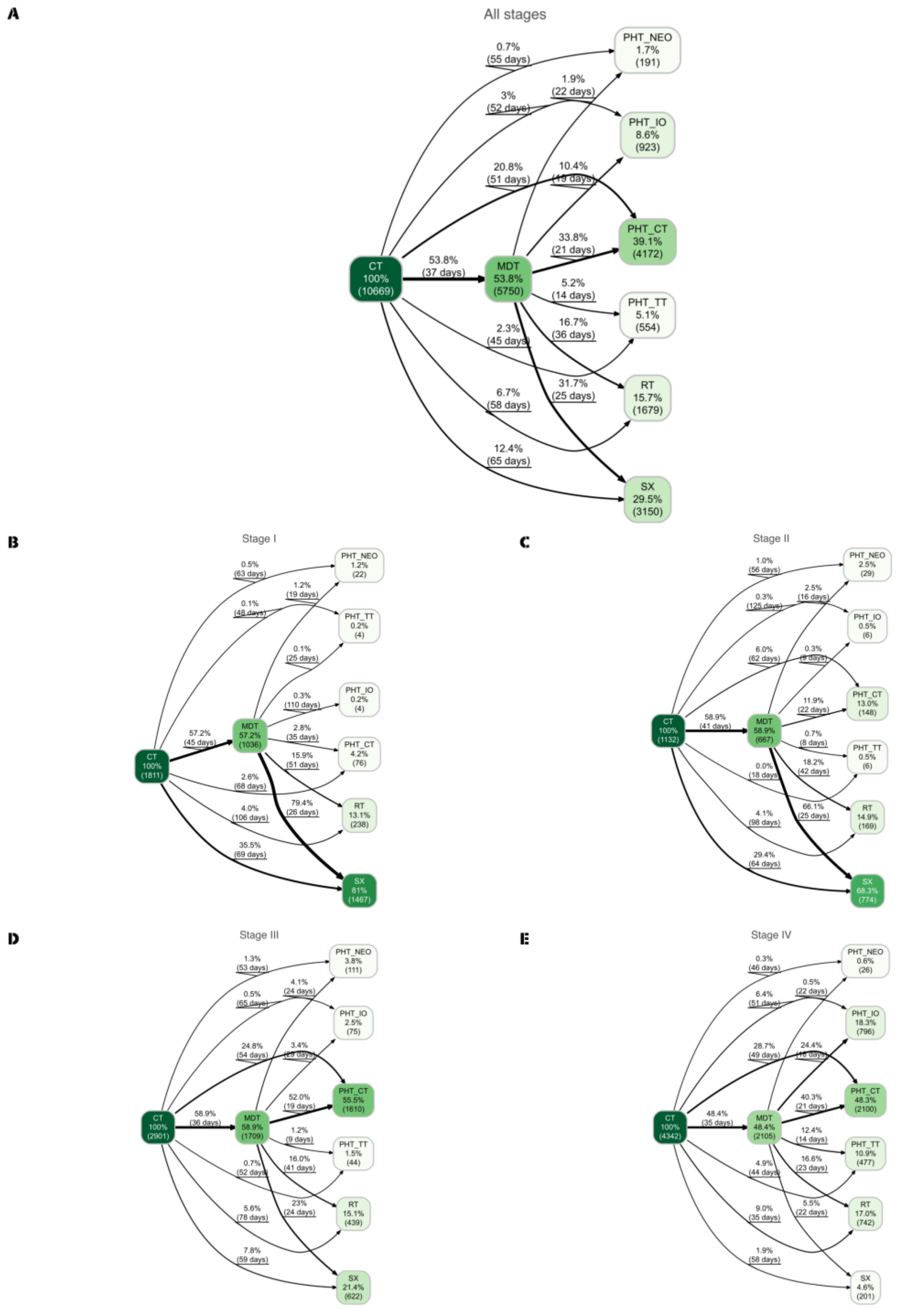


Patient pathway trajectories following the last (PET) CT in lung cancer patients. (A) The pathways illustrate the progression of all treated patients to multidisciplinary team (MDT) discussion and first-line therapy (FLT), divided into surgery (SX), radiotherapy (RT), and pharmacotherapy (PHT), which includes chemotherapy (PHT_CT), targeted therapy

(PHT_TT), and immunotherapy (PHT_IO). (B-E) The pathways illustrate treatment types stratified by disease stage.

Figure S2 Multidisciplinary team (MDT) review in the verified treated cohort with CT over time and by region

A

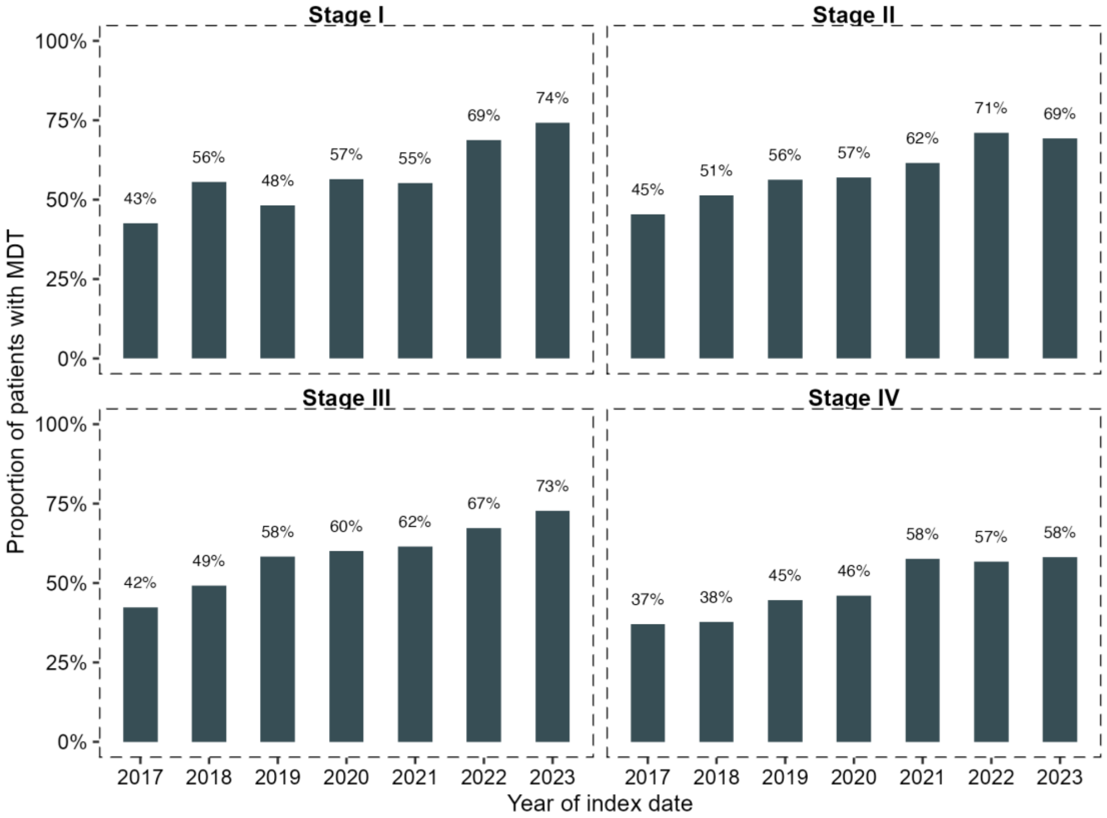


B

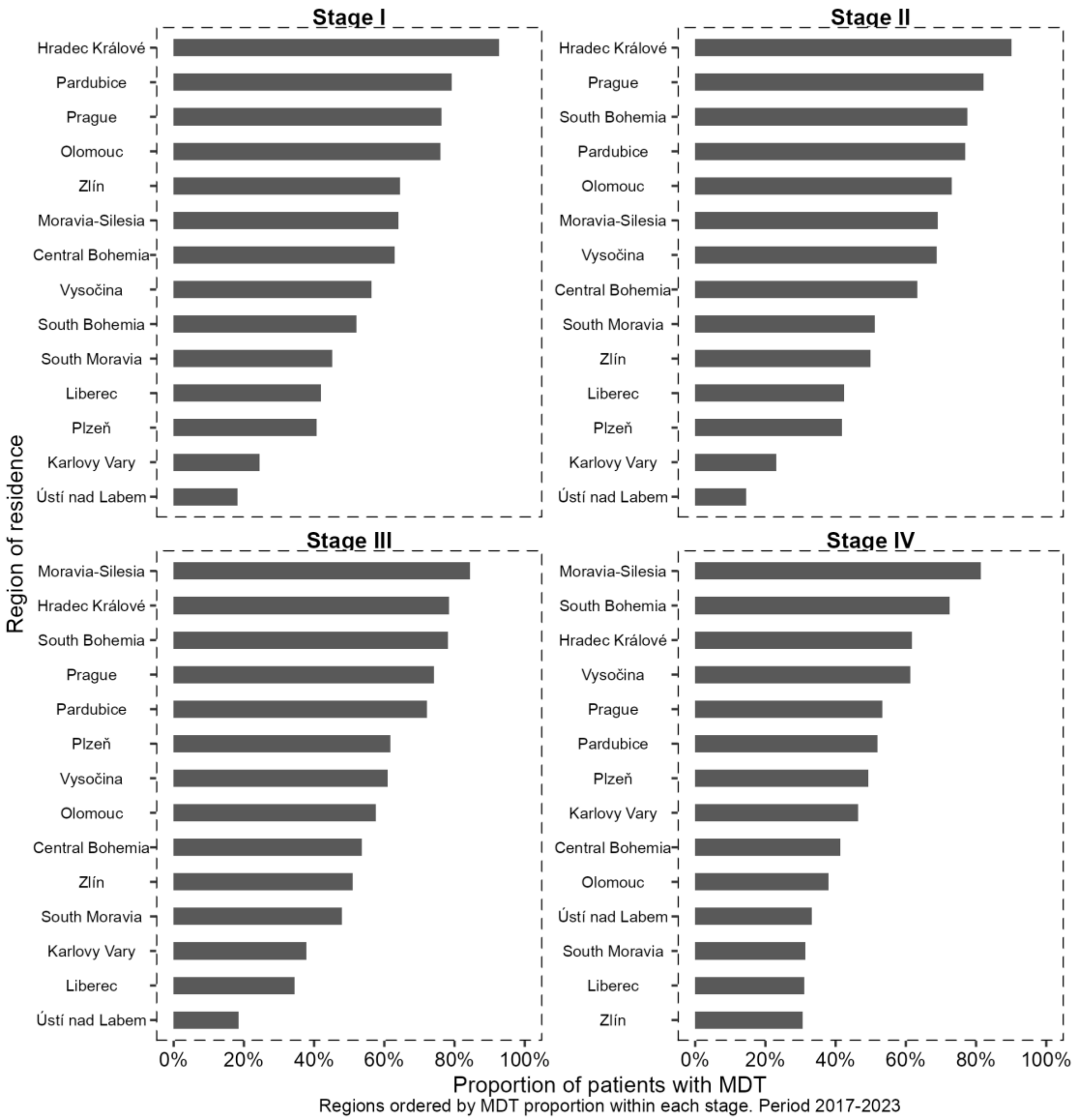


(A) MDT review by disease stage over time.
(B) MDT review by region of residence.

Table S2 Baseline characteristics of PD-L1-tested and non-tested patients (stages III–IV, 2021–2023)

Baseline characteristics by PD-L1 testing status (stages III-IV, 2021-2023)

| **Characteristic** | **Overall, N=3,151** | **Not tested, N=1,060 | ** **Tested, N=2,091** |
|---|---|---|---|
| **Sex** | | | |
| Male | 1,958 (62%) | 657 (62%) | 1,301 (62%) |
| Female | 1,193 (38%) | 403 (38%) | 790 (38%) |

| | | | |
|---|---|---|---|
| **Age** | | | |
| Mean (SD) | 69 (9) | 68 (9) | 69 (8) |
| Median (Q1, Q3) | 70 (64, 75) | 70 (64, 74) | 70 (65, 75) |
| **Stage** | | | |
| Stage III | 1,229 (39%) | 429 (40%) | 800 (38%) |
| Stage IV | 1,922 (61%) | 631 (60%) | 1,291 (62%) |
| **Year** | | | |
| 2021 | 1,052 (33%) | 391 (37%) | 661 (32%) |
| 2022 | 1,095 (35%) | 349 (33%) | 746 (36%) |
| 2023 | 1,004 (32%) | 320 (30%) | 684 (33%) |
| **FLT in COC** | 2,817 (89%) | 950 (90%) | 1,867 (89%) |
| **Region** | | | |
| Central Bohemia | 403 (13%) | 84 (7.9%) | 319 (15%) |
| Hradec Králové | 179 (5.7%) | 45 (4.2%) | 134 (6.4%) |
| Karlovy Vary | 72 (2.3%) | 49 (4.6%) | 23 (1.1%) |
| Liberec | 180 (5.7%) | 40 (3.8%) | 140 (6.7%) |
| Moravia-Silesia | 319 (10%) | 163 (15%) | 156 (7.5%) |
| Olomouc | 218 (6.9%) | 105 (9.9%) | 113 (5.4%) |
| Pardubice | 129 (4.1%) | 30 (2.8%) | 99 (4.7%) |
| Plzeň | 176 (5.6%) | 168 (16%) | 8 (0.4%) |
| Prague | 354 (11%) | 69 (6.5%) | 285 (14%) |
| South Bohemia | 208 (6.6%) | 57 (5.4%) | 151 (7.2%) |
| South Moravia | 363 (12%) | 113 (11%) | 250 (12%) |
| Ústí nad Labem | 281 (8.9%) | 49 (4.6%) | 232 (11%) |
| Vysočina | 140 (4.4%) | 22 (2.1%) | 118 (5.6%) |
| Zlín | 129 (4.1%) | 66 (6.2%) | 63 (3.0%) |

Table S3 Time to treatment initiation by disease stage and type of first-line therapy (FLT)

(A) Time to treatment from CT to first-line therapy (FLT).

Time to treatment by stage and FLT type (from CT), 2017-2023

| Stage | FLT type | N | Time to treatment, days | | |
|---|---|---|---|---|---|
| | | | Mean | 80th percentile | Median |

| | | | | | |
|---|---|---|---|---|---|
| Stage I | PHT | 106 | 128 | 126 | 72 |
| Stage I | RT | 238 | 137 | 170 | 118 |
| Stage I | SX | 1 467 | 86 | 115 | 71 |
| Stage II | PHT | 189 | 91 | 113 | 64 |
| Stage II | RT | 169 | 125 | 147 | 99 |
| Stage II | SX | 774 | 79 | 104 | 68 |
| Stage III | PHT | 1 840 | 74 | 96 | 60 |
| Stage III | RT | 439 | 100 | 133 | 81 |
| Stage III | SX | 622 | 78 | 103 | 66 |
| Stage IV | PHT | 3 399 | 71 | 90 | 55 |
| Stage IV | RT | 742 | 62 | 86 | 45 |
| Stage IV | SX | 201 | 73 | 99 | 64 |
| Stage X | PHT | 58 | 61 | 69 | 48 |
| Stage X | RT | 16 | 99 | 119 | 88 |
| Stage X | SX | 27 | 78 | 106 | 61 |
| Stage Y | PHT | 248 | 80 | 101 | 64 |
| Stage Y | RT | 75 | 103 | 155 | 78 |
| Stage Y | SX | 59 | 80 | 106 | 72 |

(B) Relative numbers of patients treated within 4-, 6-, and 8-week intervals across different years.

Time to treatment in verified cohort by stage and year

| **Stage** | **Time to treatment (from CT)** | **Year of index date** | | | | | | |
|---|---|---|---|---|---|---|---|---|
| | | **2017** | **2018** | **2019** | **2020** | **2021** | **2022** | **2023** |
| Stage I | <4 weeks | 4.0% | 4.6% | 4.4% | 5.2% | 2.1% | 3.1% | 1.5% |
| Stage I | <6 weeks | 15.4% | 15.3% | 12.3% | 15.7% | 13.3% | 10.8% | 6.5% |
| Stage I | <8 weeks | 31.2% | 26.3% | 22.5% | 28.3% | 27.8% | 22.0% | 19.2% |
| Stage I | ≥8 weeks | 68.8% | 73.7% | 77.5% | 71.7% | 72.2% | 78.0% | 80.8% |
| Stage II | <4 weeks | 5.3% | 4.5% | 2.8% | 6.4% | 3.8% | 3.6% | 3.3% |
| Stage | <6 weeks | 21.3% | 19.5% | 13.3% | 19.2% | 11.9% | 10.8% | 10.7% |

| | | | | | | | | |
|---|---|---|---|---|---|---|---|---|
| II | | | | | | | | |
| Stage II | <8 weeks | 40.0% | 37.0% | 24.9% | 32.0% | 30.2% | 26.5% | 26.0% |
| Stage II | ≥8 weeks | 60.0% | 63.0% | 75.1% | 68.0% | 69.8% | 73.5% | 74.0% |
| Stage III | <4 weeks | 10.8% | 8.1% | 6.1% | 6.6% | 7.4% | 5.2% | 5.5% |
| Stage III | <6 weeks | 32.0% | 25.3% | 21.2% | 24.1% | 23.3% | 16.4% | 18.5% |
| Stage III | <8 weeks | 51.0% | 39.9% | 38.2% | 43.6% | 43.4% | 34.4% | 36.8% |
| Stage III | ≥8 weeks | 49.0% | 60.1% | 61.8% | 56.4% | 56.6% | 65.6% | 63.2% |
| Stage IV | <4 weeks | 16.2% | 17.7% | 14.8% | 15.3% | 12.7% | 12.9% | 10.8% |
| Stage IV | <6 weeks | 37.0% | 35.6% | 36.8% | 35.2% | 31.2% | 31.0% | 29.1% |
| Stage IV | <8 weeks | 56.8% | 53.8% | 52.7% | 54.7% | 49.1% | 49.0% | 49.3% |
| Stage IV | ≥8 weeks | 43.2% | 46.2% | 47.3% | 45.3% | 50.9% | 51.0% | 50.7% |
| Stage X | <4 weeks | 22.2% | 7.5% | 50.0% | - | 50.0% | - | 50.0% |
| Stage X | <6 weeks | 33.3% | 28.3% | 100.0% | - | - | 33.3% | 100.0% |
| Stage X | <8 weeks | 41.7% | 45.3% | - | - | 100.0% | 66.7% | - |
| Stage X | ≥8 weeks | 58.3% | 54.7% | - | 100.0% | - | 33.3% | - |
| Stage Y | <4 weeks | 7.8% | 3.8% | 4.2% | 9.5% | 5.9% | 5.6% | 5.1% |
| Stage Y | <6 weeks | 17.6% | 20.8% | 18.1% | 21.4% | 13.7% | 18.5% | 13.6% |
| Stage Y | <8 weeks | 45.1% | 39.6% | 38.9% | 35.7% | 25.5% | 33.3% | 28.8% |
| Stage Y | ≥8 weeks | 54.9% | 60.4% | 61.1% | 64.3% | 74.5% | 66.7% | 71.2% |

Figure S3 Time to MDT and from MDT to treatment initiation in verified treated cohort

A

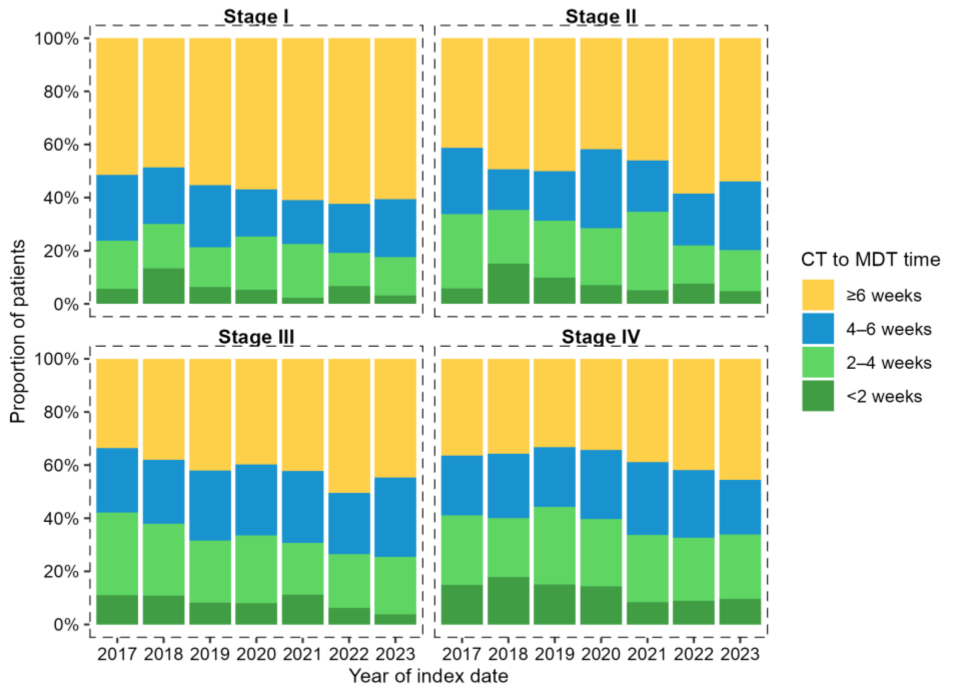


B

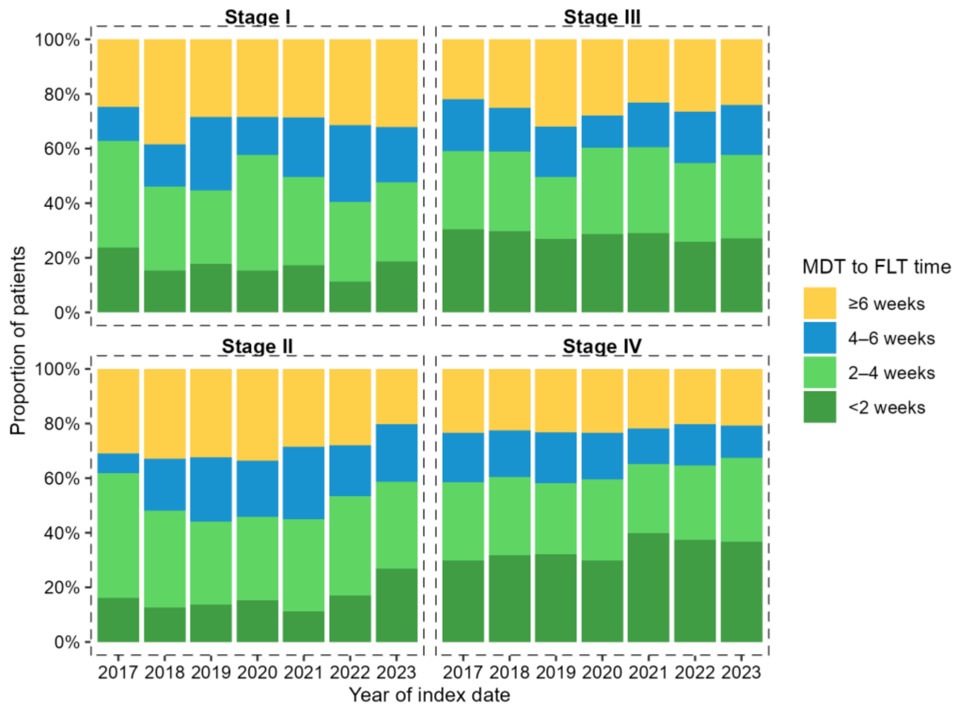


(A) Time from (PET) CT to multidisciplinary team (MDT) discussion.

(B) Time from MDT to first-line therapy (FLT).

Figure S4 Time to treatment initiation by disease stage and region of residence

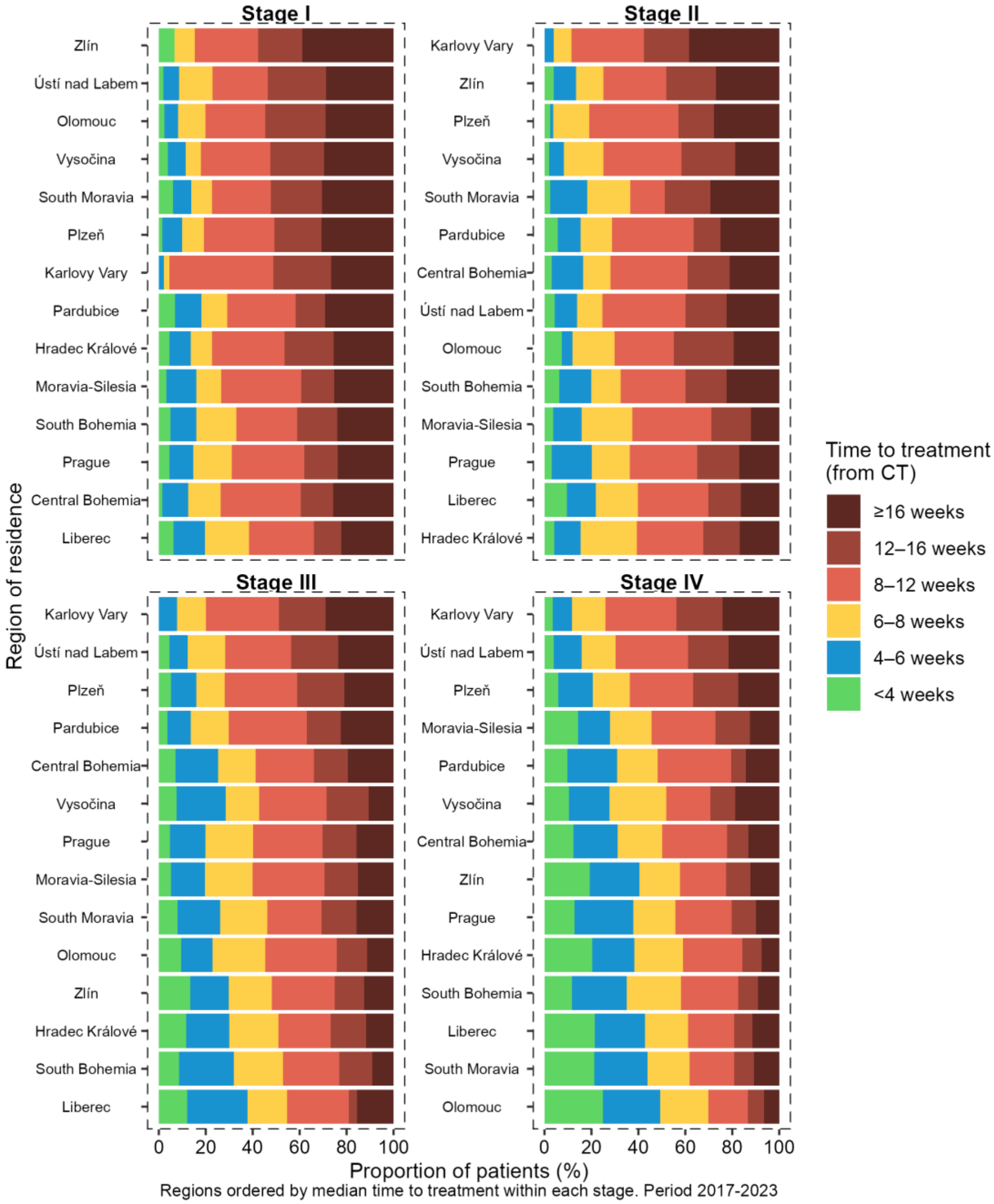

Figure S5 First-line therapy (FLT) in Complex Oncology Centres (COCs) for treated patients by region of residence and disease stage

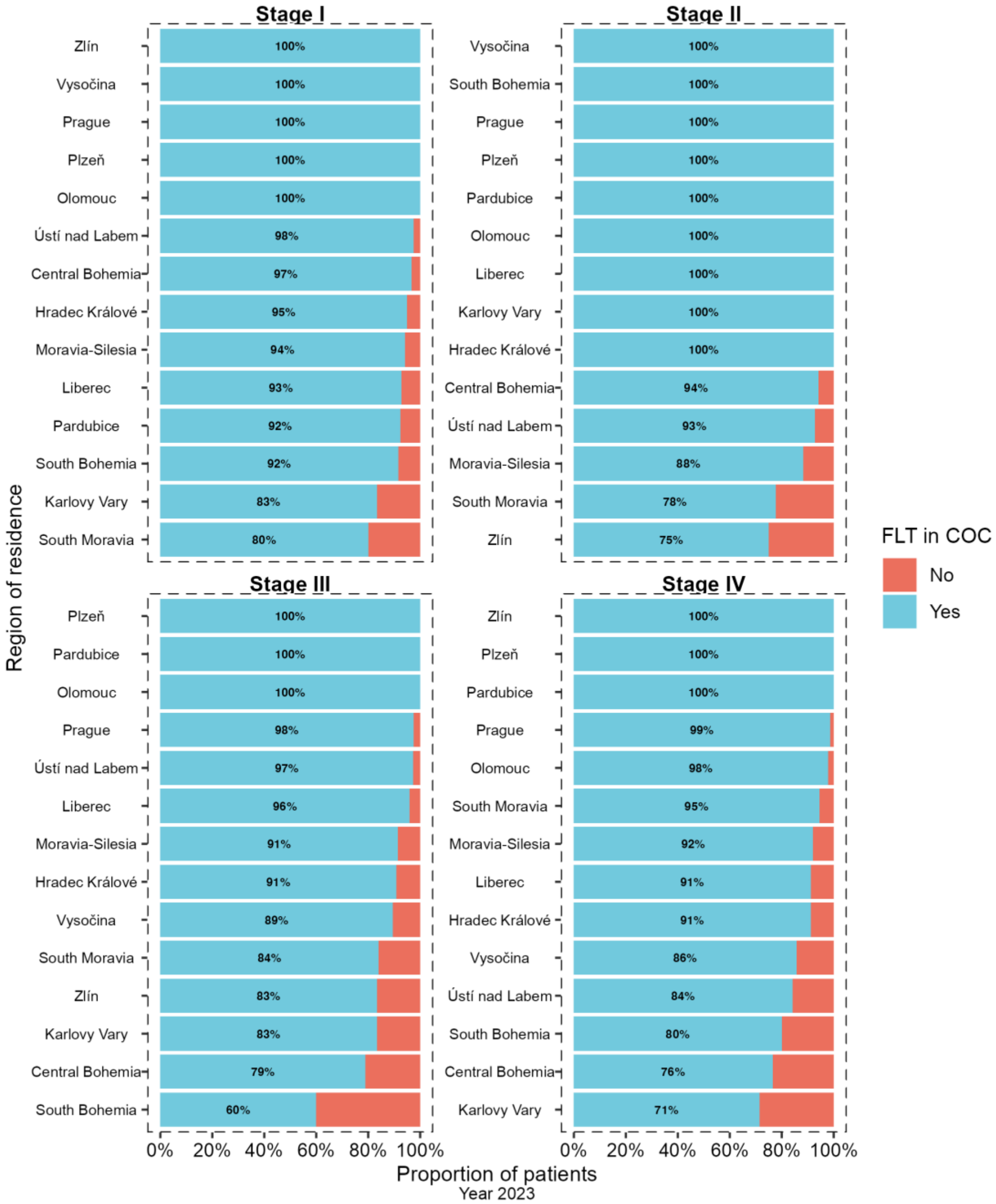
